# *AnmO*_2_*l*: An open-source pulse-dose oxygen conserving device for the COVID-19 crisis

**DOI:** 10.1101/2021.05.19.21257477

**Authors:** Honquan Li, Deepak Krishnamurthy, Anesta Kothari, Ethan Li, Michael Lipnick, David Gaba, Ruth Fanning, Manu Prakash

**Affiliations:** Electrical Engineering, Stanford University; Bioengineering, Stanford University; UCSF Center for Health Equity in Surgery and Anesthesia; Anesthesiology, Perioperative and Pain Medicine, Stanford University; Woods Institute for the Environment; CZI BioHub Investigator

## Abstract

The surge of COVID-19 cases in India and around the world has resulted in acute oxygen shortage. Oxygen therapy through nasal cannula with flow rates of 1-6 L/min is an effective treatment for many COVID-19 patients in non-critical conditions and may help prevent disease progression. The same treatment is also used in post-acute care for recovering COVID-19 patients. Here we describe a simple, open source, and rapidly manufacturable oxygen conservation device for use with dual-port nasal cannula that can extend the life of current oxygen supply by almost two to three times, which we hope will help towards coping with the ongoing crisis.

## 1. Introduction

The Indian sub-continent and parts of south-east Asia are currently facing a devastating second wave of the COVID-19 pandemic. Oxygen shortage is currently an acute issue in many major cities and towns where infections have spiked. At the time of writing (May 17, 2021), estimates based on WHO data put the current oxygen demand in India at 15.8 million cubic meters or 2.2 million oxygen cylinders per day [1]. These numbers which are based on testing data are also likely underestimates. Challenges in both production and supply-chain management and distribution mean that the country is struggling to distribute the life-saving supply where it is needed the most [2]. The problem is only more severe as the disease further spreads in the rural areas, which have two-thirds of India’s population but have less infrastructure and are associated with more transportation delays. Worryingly, other countries in the Indian sub-continent like Nepal have also started facing acute oxygen shortages mirroring the situation in India [3].

Among symptomatic COVID-19 patients, while most only develop mild (40%) and moderate (40%) disease, around 15% develop severe disease requiring oxygen support, whereas 5% have critical disease with complications requiring intensive care [4]. For those patients who require supplemental oxygen, in the course of their clinical management, delivery of oxygen usually starts and stops with nasal cannula at flow rates up to 6 l/min. *Oxygen delivered during patient exhalation is mostly vented to the air and wasted, providing an opportunity for oxygen conservation by shutting the flow when the patient is exhaling*. Towards this, some completely passive solutions have been developed including Oxymizer [5, 6]. However, due to the limited reservoir size and fluid dynamic parameters which these devices are designed for, the effect of oxygen saving decreases significantly as the flow rate is increased (3:1 at 0.5 L/min, 1.7:1 at 2 L/min, 1.4:1 at 4 L/min and 1.3:1 at 5 L/min) [7] making these devices useful primarily in oxygen therapy for mild, stable respiartory failure (e.g. chronic obstructive pulmonary disease (COPD). Other more effective oxygen conservation devices have been developed in the 1990’s and early 2000’s, but now have very limited or no availability.

Here we describe an open source implementation of a simple yet effective oxygen conservation device which makes use of readily available parts and can be rapidly manufactured in centralized or distributed approaches by reputed manufacturers. While we pursue larger scale clinical evaluation, through this rapid communication we would like to share the basic design and initial testing of the device, obtain feedback, and connect with clinicians as well as interested design and manufacturing partners to be able to bring this solution to patients and support oxygen delivery and conservation efforts in South-Asia.

## 2. Results

There is a large design space for oxygen conservation devices (electronic vs pneumatic, variable pulse length^1^ vs variable flow rate^2^ etc.) [8,9]. We choose the electronic route so that manufacturers can rely on mature, off-the-shelf components that are already certified for performance and reliability. To further settle on a design, we asked ourselves what is the simplest design that is safe, effective, easy to use, and easy to manufacture rapidly and at scale. This design philosophy leads to the following requirements:

1. The device should at minimum be able to interface with an oxygen cylinder through the most common interfaces - standard oxygen regulators which (1) reduce the supply to 50 psi and (2) can be used to set continuous flow rate. The same principles might also be applicable to delivery of oxygen via manifolds.
2. The overall system (the device + parts connected to the device) should ideally only have one adjustable control parameter for setting the flow profile, and the flow profile should be associated with an equivalent continuous flow rate that healthcare providers can easily read out. Since the device works with an already-existing oxygen delivery system (oxygen tank and nasal cannula) or manifold, other parameters should still be adjusted using already-existing control knobs to avoid the need to control two redundant interfaces.
3. The device should provide continuous flow with flow rates that are not much higher than the set equivalent continuous flow rate when the device is disconnected from power or when the device fails for any other reason (fail-safe by fail-open approach).
4. It should be simple to bypass the system or introduce it in a system on an on-demand basis when oxygen needs change dramatically.

These requirements dictates that we should use a variable flow rate approach and narrows the design down to one that uses an on/off normally-open valve, where the continuous flow rate is the flow rate set on the oxygen regulator. Such a design, that is used in our current implementation, is shown in Figure 1.

**Fig. 1.**
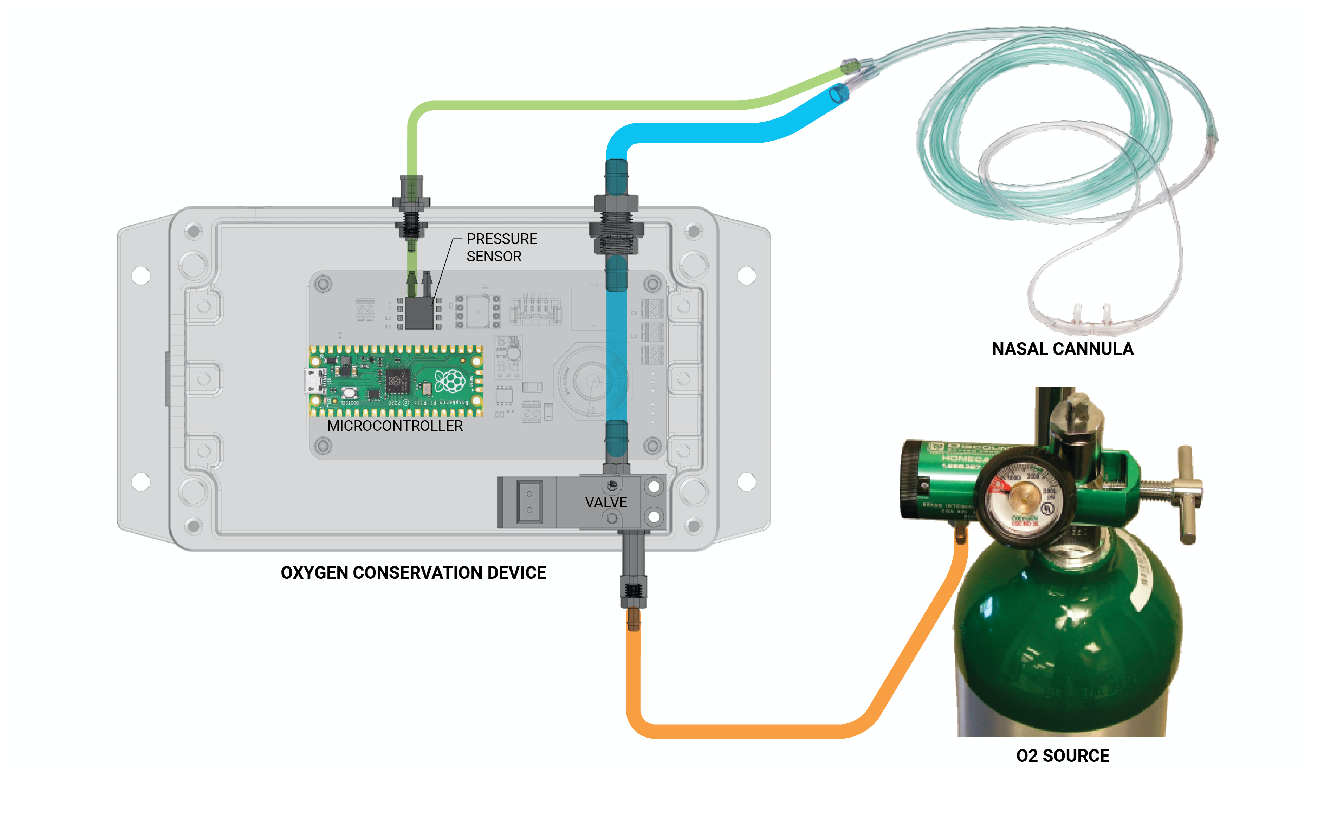
Simplified schematic and piping diagram of the proposed device with major parts labelled above, including source of oxygen (either a cylinder or oxygen manifold), pulse-dose device with a fail-open ON/OFF valve, a pressure sensor and a micro-controller connected to dual probe nasal cannula and a re-chargeable battery that can be swapped. The device is encapsulated in a water proof enclosure and can sit on the bed side accommodating various length of the nasal cannula.

To keep it simple and modular, we choose an enclosed design that does not have a built-in battery but exposes a USB micro-B or USB-C connection for powering the device, either through a 5V wall adapter or a battery pack. In its simplest design, the exposed USB port is the only human-machine interface: the user would plug in power to turn on the device, enabling oxygen conservation, and disconnect power to turn off the device for receiving continuous flow (effectively bypassing the device with an valve-open state). For enhanced user-machine interaction, an ON/OFF switch and LED indicator has been added to indicate the power and operation state of the device. Further more, we have included a medical alarm on the PCB, which may be used to alarm the health care provider, for example, of (prolonged) high respiratory rate, an indicator of distress and requirement of increased flow rate or further escalated care. In case the medical alarm is included, an alarm silencing button can also be included. A photo of the current prototype and the CAD rendering of the complete design can be found in Supplementary Figure 3 and Supplementary Figure 4

For the current iteration, we chose to have a dedicated pressure port for use with dual limb nasal cannula^3^, so that the pressure measurement is not affected by the presence of flow, which makes the software implementation of precise valve on/off timing straightforward and flexible. For example, threshold values can be set so that flow delivery starts near the end of the exhalation (before inhalation starts) and stops before the end of inhalation for higher oxygen delivery and conservation efficiency.

As an initial test, a volunteer trialed the device (with the O2 limb cut as we’re waiting for IRB approval) and the recorded pressure and flow waveform is shown in Figure 2. We note that the spike at the beginning of the flow delivery is reduced when an intact nasal cannula is connected to the device (Supplementary Figure 5). We also confirmed that the initial peak is due to the 50 psi reservoir largely formed by the tubing connecting the oxygen regulator and the device (Supplementary Figure 6). Such peaks, with limited magnitude (less than 10 l/min in all cases) should not result in user discomfort and will increase oxygen delivery efficiency, according to prior studies [8, 9]. Lastly, we conducted initial tests of mouth breathing and coughing, with satisfactory result (Supplementary Figure 7).

**Fig. 2.**
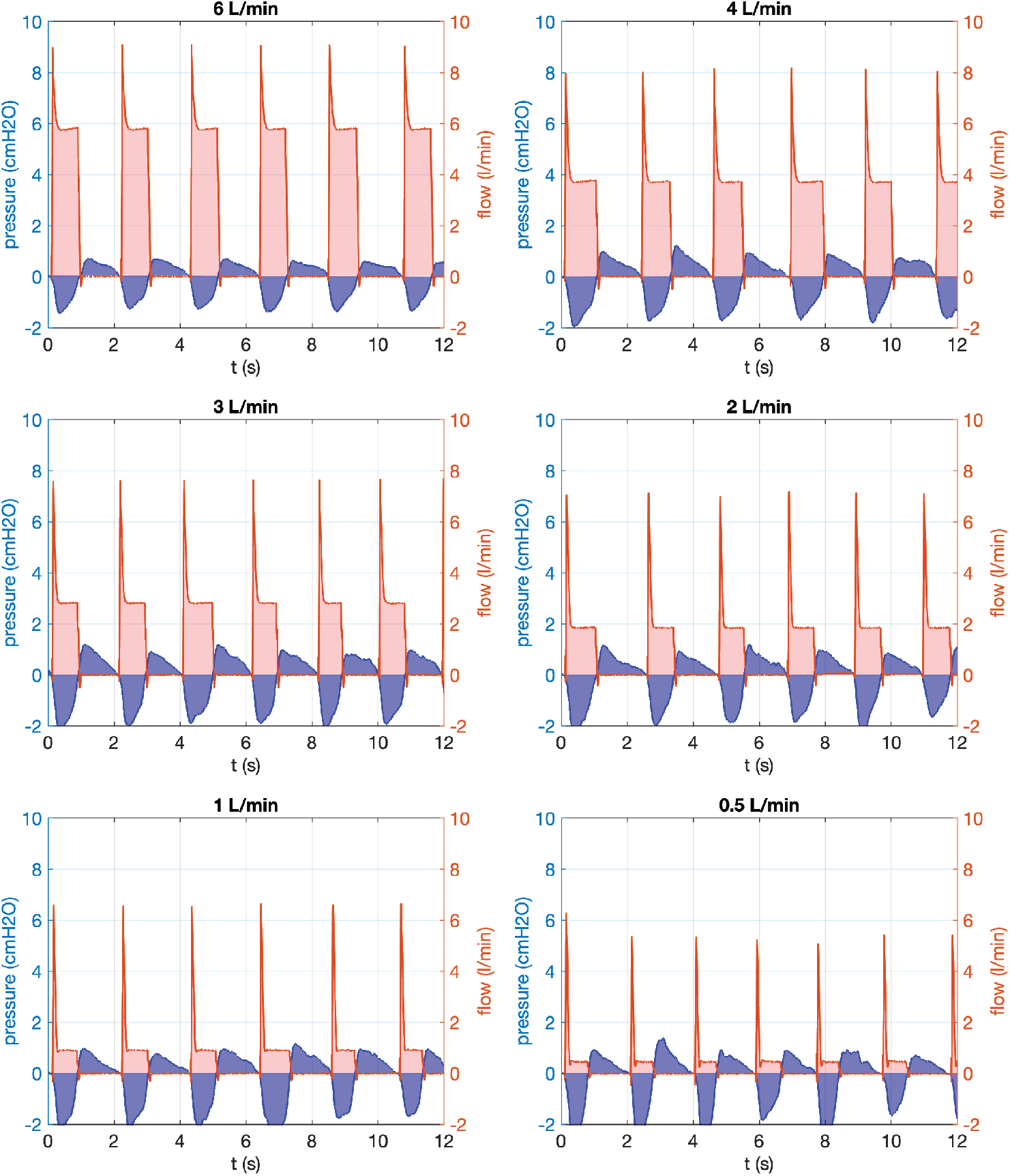
Sample flow-rate and pressure waveforms from the prototype device at different flow-rates. The flow-rates are set by the flow-regulator attached to the oxygen cylinder and are not controlled by the device.

## 3. Discussion

In this rapid communication we presented the design, implementation and initial testing of a new rapid manufacturable oxygen conservation device that is developed with the goal of helping mitigate the adverse effects of oxygen shortages.

Comparison of the measured waveform with validated devices [8, 9] suggests that satisfactory performance may be expected, which should be confirmed by further testing and clinical evaluations.

Right now the device is only indicated for use with nasal cannula. While we have confirmed that trigger also works with simple face masks that traditionally require 5-10 l/min of flow for oxygen enrichment and CO2 washout (Supplementary Figure 8), whether there are trigger timing schemes that allow the use of the device with simple face masks remains to be studied.

Lastly, additional features may be built on top of the current implementation. Examples include logging of respiratory rate and other respiratory parameters, display of trend(s), Bluetooth interface with SpO2 sensor for alarms, Bluetooth interface with a device hub for remote monitoring.

## Materials and Methods

### Implementation

The device is controlled by a Raspberry Pi Pico microcontroller, and uses temperature compensated differential pressure sensor with either SPI or I2C interface. The microcontroller is programmed by the Arduino IDE. A Clippard 15 mm normally open solenoid valve is controlled through a PWM valve driver IC that implements “hit and hold” for reduced power consumption and heat generation. A custom PCB is designed in KiCAD, fabricated, and used in the prototypes. The firmware of the device is hosted on Github at https://github.com/prakashlab/pulse-dose-device. In the first prototype, the enclosure is 3D printed. 1/8” ID, 1/4” OD AdvantaPure APHP High Pressure Unreinforced Silicone Tubing is used to connect the device and the oxygen regulator. Associated files can be downloaded at project website at www.anmo2l.org.

### Initial tests and characterizations

For testing and characterizations, a Sensirion SFM3019 flow sensor is directly connected to microcontroller, through a reserved connector on the PCB. Pressure and flow are measured every 5 ms and transmitted through Serial Interface over native USB connection to a computer for recording. In the initial device test, a volunteer wears the nasal cannula (Teleflex 2845) but the O2 limb is cut so no oxygen is delivered to the test subject. To repeatably measure the flow profile, the firmware is modified to generate software trigger. During this test, an intact nasal cannula is connected.

#### Nose vs mouth breathing test

The proposed device should be sensitive enough to detect patient breath over a wide range of conditions including in nose vs mouth breathers. To test this sensitivity the device was connected to a dual-lumen nasal cannula worn by a volunteer with no oxygen cylinder connected, and the pressure waveforms recorded via the on-board pressure sensor on the device. The volunteer was asked to breathe normally through the nose for a few breaths and then asked to breathe through the mouth. Consciously breathing through the mouth lowered the amplitude of the negative pressure peaks by around 3-4x, however, still resulting in clearly resolved peaks (Supplementary Figure 7A). These peaks were detected robustly by the device and triggered a breath. The triggering could only be confused by completely blocking the nose by pinching with fingers and isolating the nasal prongs from the mouth.

#### Cough test

On the other end of the spectrum we tested how the device responded during patient coughs. The setup was the same as that of the nose-vs-mouth breathing test described above. The volunteer was asked to simulate coughing and the pressure waveforms recorded. The device was able to initiate a breath in the inspuratory periods that punctuate the coughs thus performing as expected. These preliminary tests show that the device performs as expected even with single pressure threshold value.

## Data Availability

All data available at www.anmo2l.org

## Acknowledgement

We thank Dr. Bijay Acharya, M.B.B.S., M.P.H., Dr. Bhavna Seth, MD, MHS, Pulmonary Critical Care Fellow, Johns Hopkins University, Dr. Nitin Arora, MD, M.P.H, University of Alabama, Dr. Kai Kuck University of Utah, Dr. Daniel Harris, Brown University, Dr. Subir Bhaduri and Brian Daniel for clinical and technical feedback and valuable suggestions. We also thank clinicians and engineers in the IndiaCovidSOS group for valuable feedback.

## Supplementary Figures

**Fig. 3.**
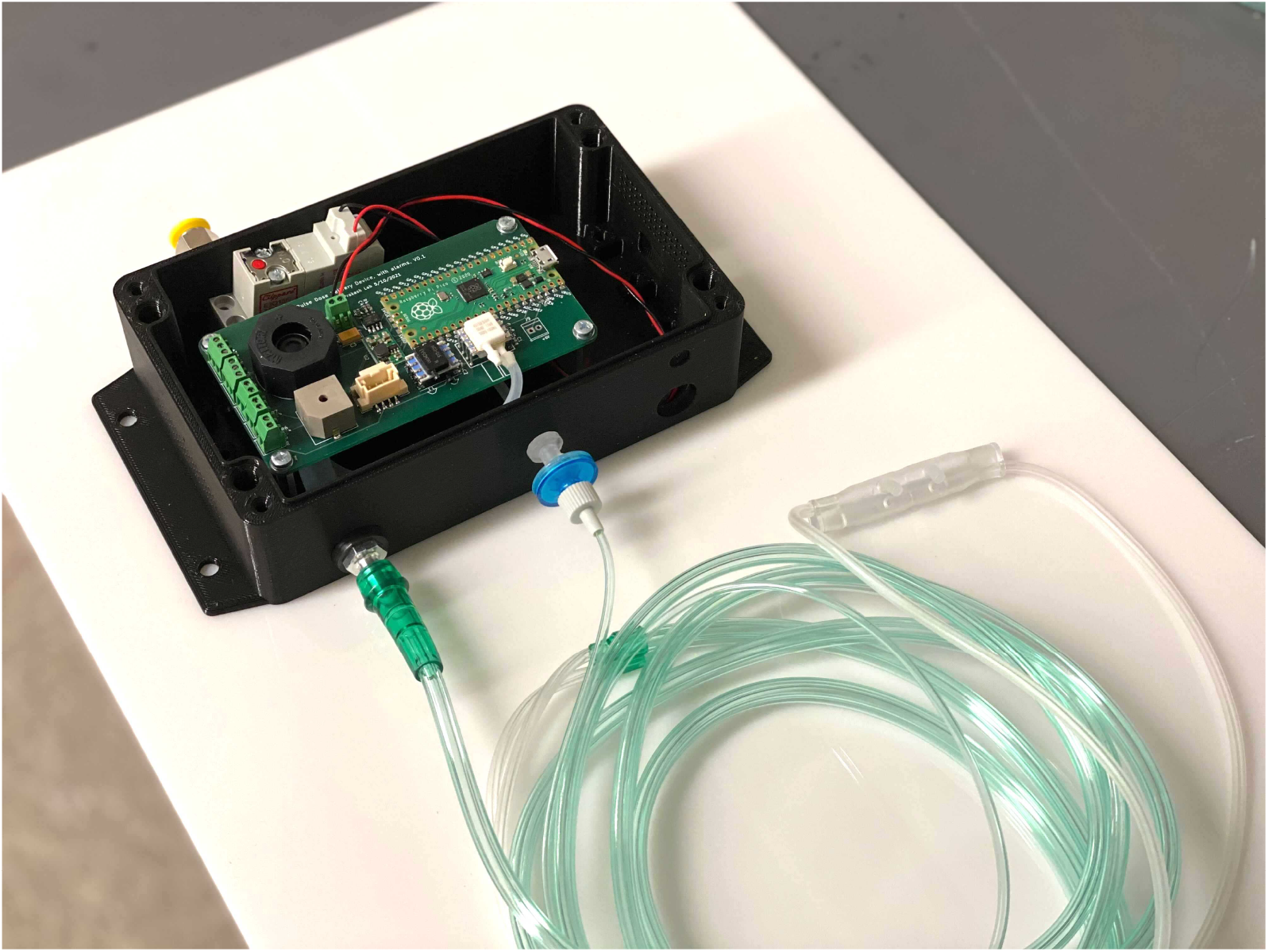
Photograph of the prototype device used for testing. The photo shows the device with the enclosure cover removed. During normal operation the cover will be closed.

**Fig. 4.**
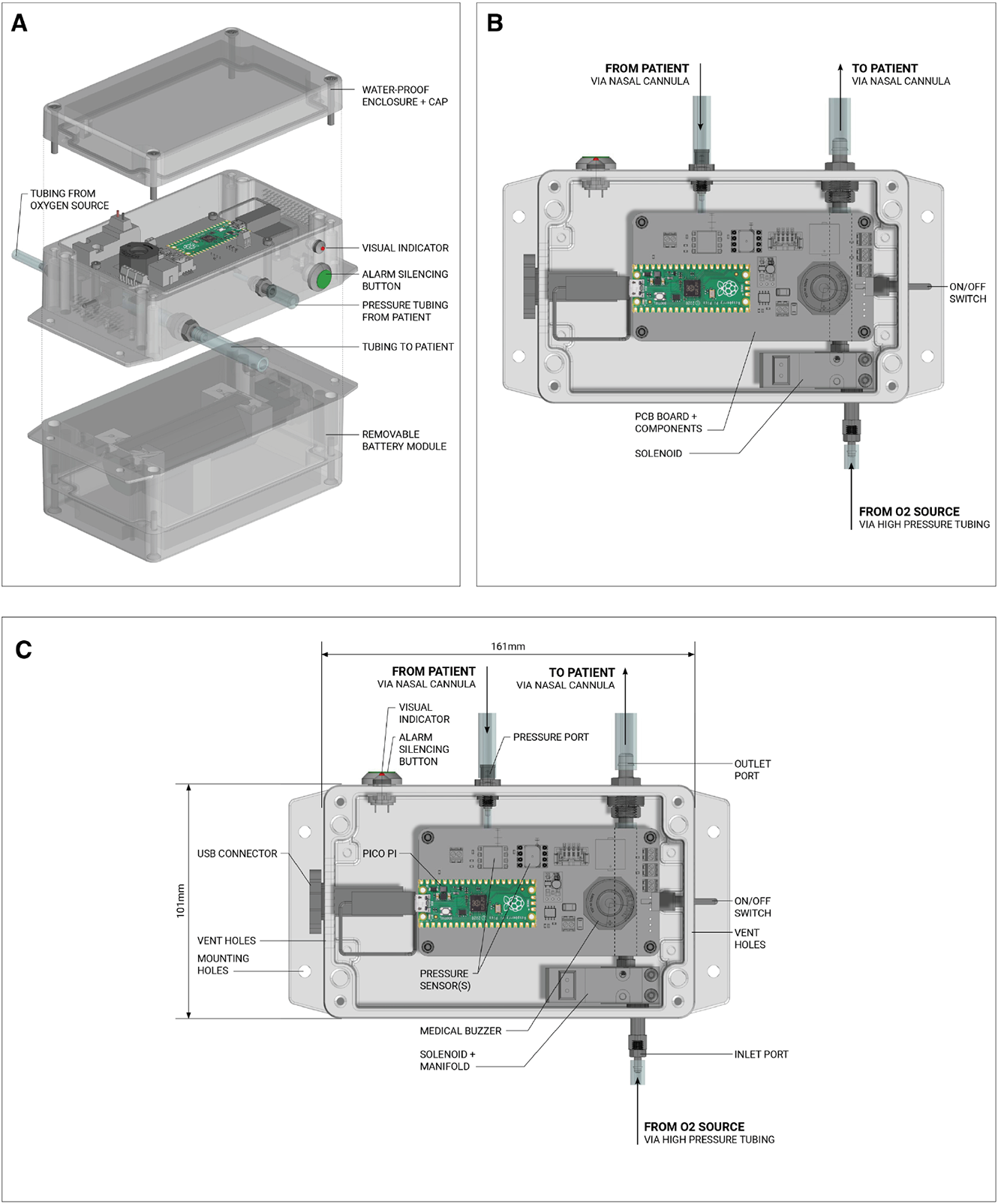
Full CAD rendering of the pulse-dose-device. (A) Showing both main enclosure and battery module. (B) and (C) plan view of the device with major parts labelled.

**Fig. 5.**
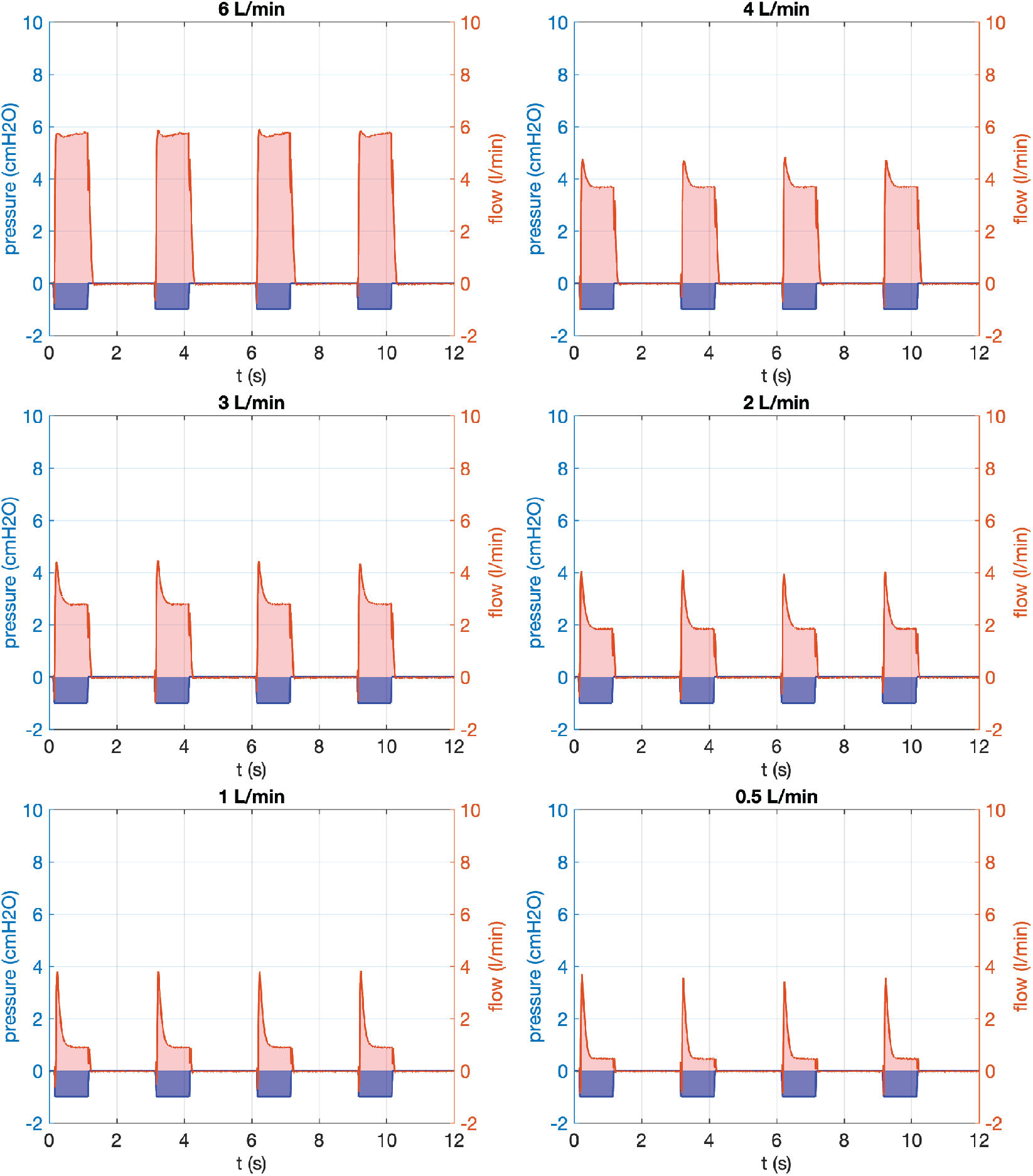
Effects of upstream tubing length. The upstream tubing is around 1 ft long. Recorded waveform with software simulated trigger.

**Fig. 6.**
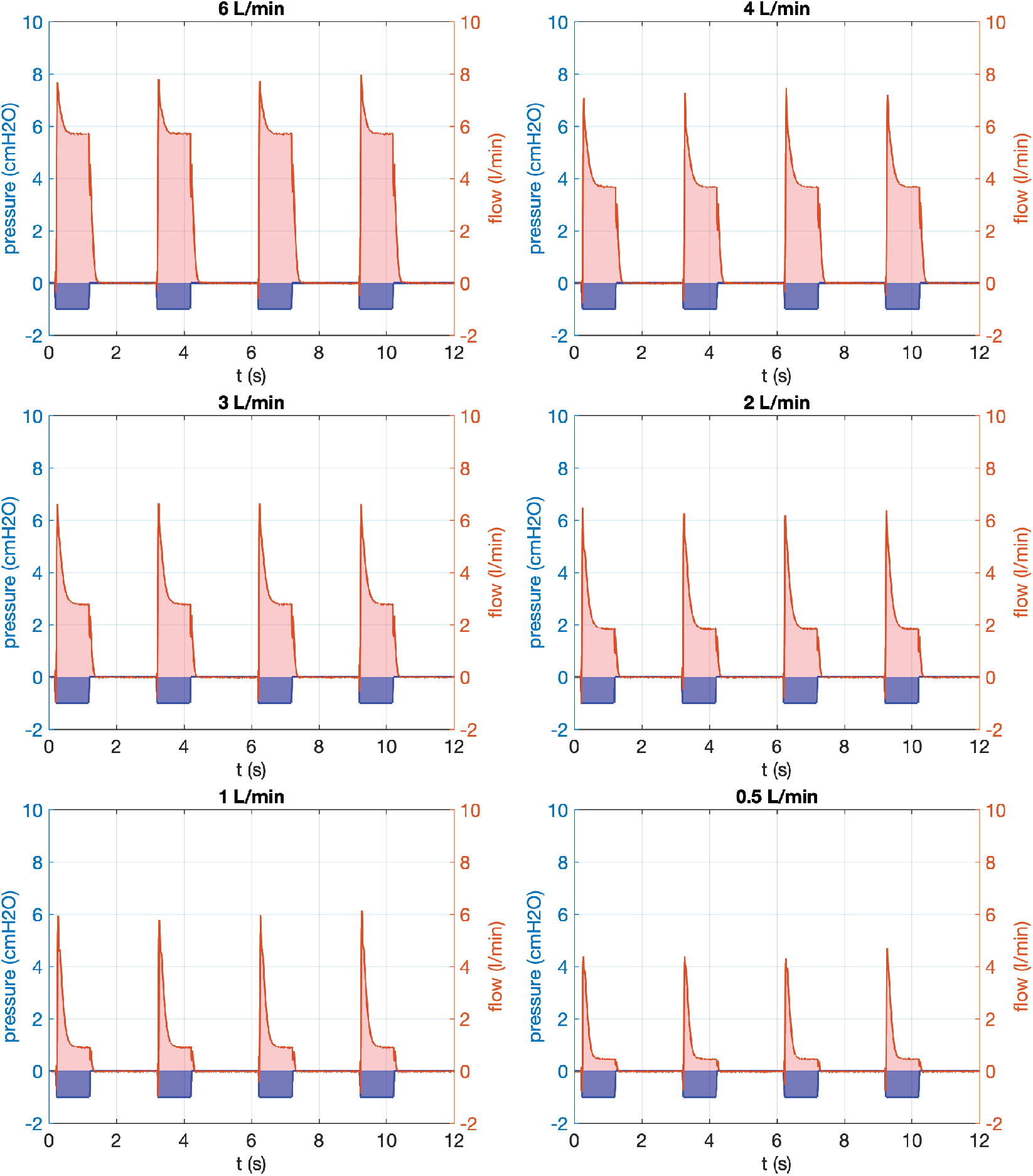
Effects of upstream tubing length. The upstream tubing is around 2 ft long. Recorded waveform with software simulated trigger.

**Fig. 7.**
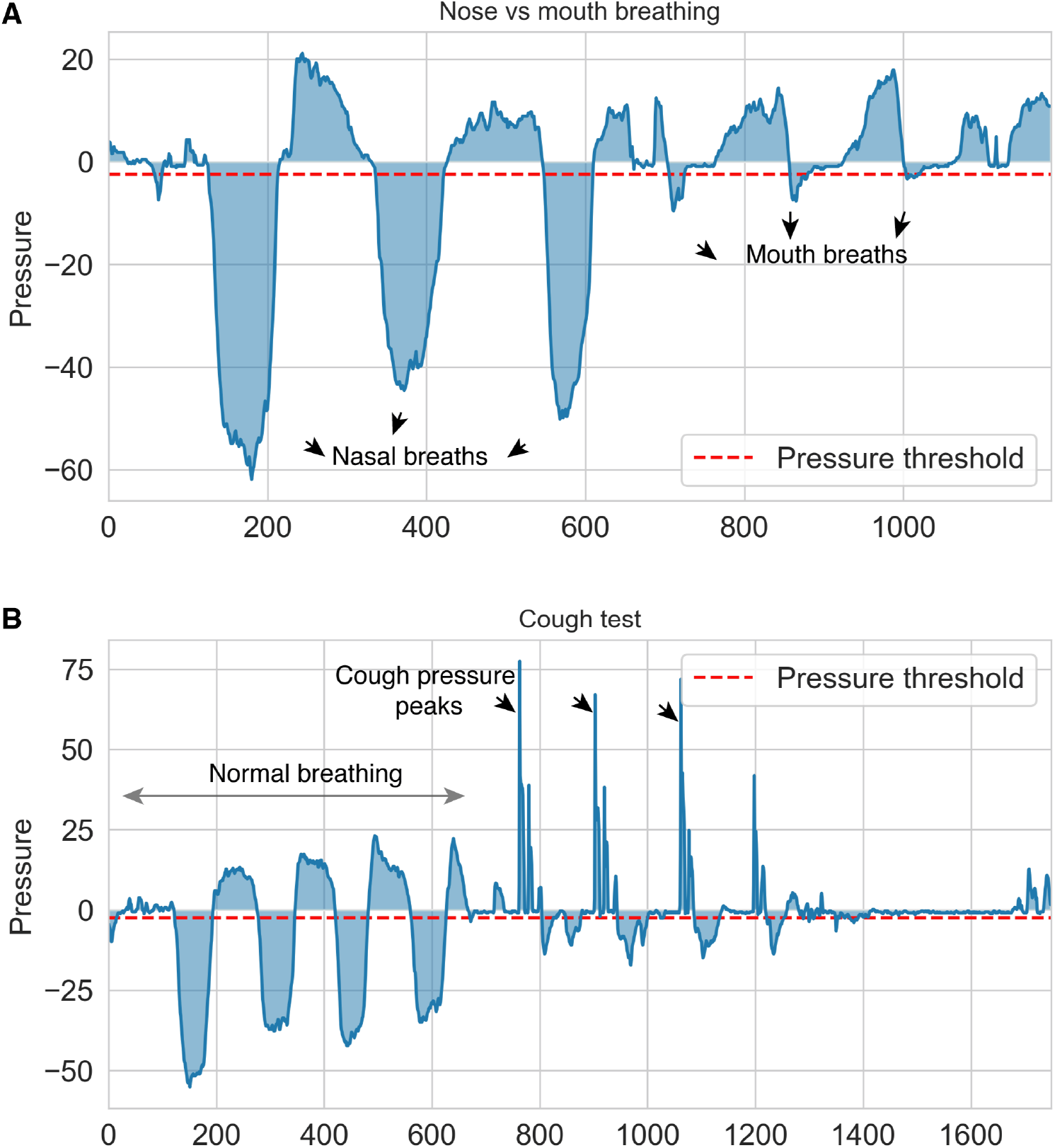
(A) Testing trigger sensitivity during nose vs mouth breathing. (B) Testing device performance during patient coughs.

**Fig. 8.**
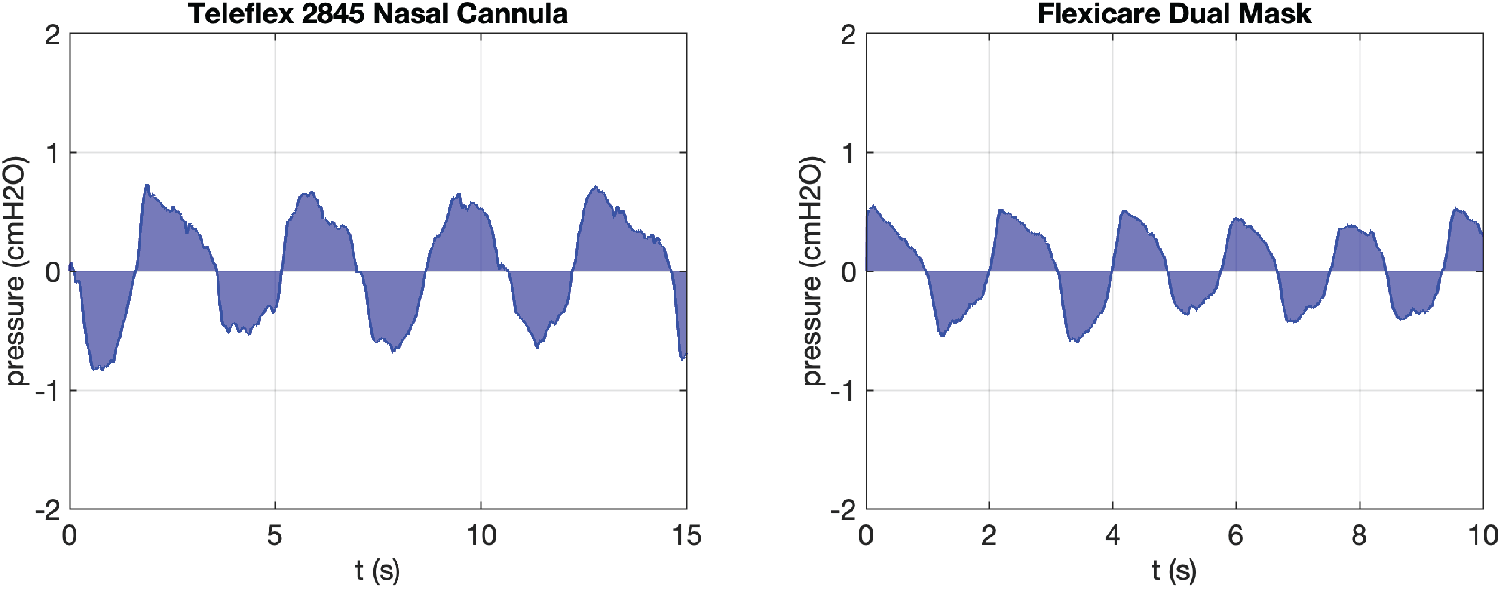
Recorded pressure waveform for nasal cannula and simple face mask.

## Footnotes

1 fixed flow rate, change the pulse length to change the equivalent continuous flow rate

2 fixed pulse length - relative to the patient breathing pattern, change the flow rate to change the equivalent continuous flow rate

3 these nasal cannulas are also standard and widely used for CO2 measurement and O2 delivery

## References

1. “COVID-19 oxygen needs tracker,” https://www.path.org/programs/market-dynamics/covid-19-oxygen-needs-tracker/.

2. Reuters, “Why india is facing an oxygen crisis as covid-19 cases mount,” Hindustan Times (2021).

3. H. Ellis-Petersen, “‘a hopeless situation’: oxygen shortage fuels nepal’s covid crisis,” The Guard. (2021).

4. W. H. Organization et al., “Covid-19 clinical management: living guidance, 25 january 2021,” Tech. rep., World Health Organization (2021).

5. B. L. T. E. P. A. Otsap, “Oxygen delivery apparatus,” (U.S. Patent 4 535 767 A, Oct. 1982).

6. E. R. Abel, “Oxygen delivery and conserving device,” (U.S. Patent 5 280 780 A, Nov. 1992).

7. C. P. Dumont and B. L. Tiep, “Using a reservoir nasal cannula in acute care,” Critical Care Nurse 22, 41–46 (2002).

8. R. W. M. R. BSM, “A bench study comparison of demand oxygen delivery systems and continuous flow oxygen,” Respir. care 44, 925 (1999).

9. V. I. Products, “Your 2007 guide to understanding oxygen conserving devices,” http://www.inspiredrc.com/images/documents/OCD2007.pdf (2007).

